# The Independence of Discrimination and Calibration in Clinical Risk Prediction: Lessons from a Multi-Timeframe Diabetes Prediction Framework

**DOI:** 10.64898/2026.02.12.26346147

**Authors:** Eoghan O’Reilly, Tomas Kurakovas

## Abstract

**Background:** Clinical risk prediction models are typically evaluated by discrimination (area under the receiver operating characteristic curve, AUC), with calibration receiving less attention. We developed a multi-timeframe diabetes prediction framework emphasizing calibration and used synthetic data validation to investigate whether good discrimination guarantees good calibration.

**Methods:** We generated 500,000 synthetic patients using published epidemiological parameters from QDiabetes-2018, FINDRISC, and the Diabetes Prevention Program. The framework comprises a discrete-time survival ensemble with isotonic calibration, producing predictions at 6, 12, 24, and 36 months with bootstrap confidence intervals. We evaluated discrimination (AUC), bin-level calibration (expected calibration error, ECE), calibration-in-the-large (observed-to-expected ratio), and clinical utility (decision curve analysis). We compared performance against QDiabetes-2018 implemented on the same synthetic cohort.

**Results:** Despite achieving excellent discrimination (AUC = 0.844, 95% CI: 0.840-0.848) and low bin-level calibration error (ECE = 0.006), the framework systematically overpredicted risk by 50%: mean predicted probability was 8.4% versus observed rate of 5.6% (observed-to-expected ratio = 0.66, 95% CI: 0.65-0.67). This miscalibration occurred despite isotonic regression on a held-out calibration set. Overprediction was present in 9 of 10 risk deciles. Risk stratification remained valid (23.5-fold separation, 95% CI: 22.8-24.3, between highest and lowest tiers), confirming that discrimination was preserved. QDiabetes-2018 achieved comparable discrimination (AUC = 0.831) with better calibration (O:E = 0.89). Decision curve analysis showed net benefit across threshold range 5-30%, though recalibration would improve clinical utility.

**Conclusions:** Good discrimination does not guarantee good calibration. Our primary finding is negative: isotonic calibration failed to produce well-calibrated predictions even on synthetic data from a single generator. This has important implications for model deployment, where distribution shift is inevitable. We recommend that prediction model studies report calibration-in-the-large alongside bin-level metrics, as ECE alone can be misleading when risk distributions are skewed. Recalibration on deployment populations will likely be necessary for any prediction model, regardless of development-phase calibration performance.

**Key Messages:** *What is already known:*

- Clinical prediction models require both discrimination (ranking patients correctly) and calibration (accurate probability estimates)
- Isotonic regression is a recommended approach for post-hoc calibration
- Expected calibration error (ECE) is commonly reported as a summary calibration metric

*What this study adds:*

- Demonstrates empirically that excellent discrimination (AUC = 0.844) can coexist with substantial miscalibration (50% overprediction)
- Shows that low ECE can be misleading when most patients fall in low-risk deciles
- Provides evidence that isotonic calibration on held-out data may not generalize even within synthetic data from one generator
- Demonstrates a discrete-time survival architecture that reduces monotonicity violations to <0.1%

*How this study might affect research, practice, or policy:*

- Prediction model studies should report calibration-in-the-large (O:E ratio) alongside ECE
- Developers should expect recalibration to be necessary when deploying to new populations
- Claims of ‘calibrated prediction’ should be viewed skeptically without comprehensive calibration assessment

## 1. Introduction

### 1.1 The Calibration Problem in Clinical Prediction

Clinical risk prediction has become central to precision medicine, enabling personalized screening, treatment selection, and shared decision-making. The predominant evaluation paradigm focuses on discrimination-the ability to rank patients by risk, typically measured by the area under the receiver operating characteristic curve (AUC). A model with AUC of 0.85 is generally considered ‘good,’ and developers often proceed to implementation based primarily on this metric.

However, clinical decision-making depends not on rank-ordering alone but on probability accuracy. When a model predicts 40% risk, clinicians and patients need confidence that approximately 40% of similar patients experience the outcome. This property-calibration-has received comparatively less attention in the machine learning literature, despite being essential for clinical utility.

Van Calster and colleagues demonstrated that even models with excellent discrimination can be clinically harmful if miscalibrated, potentially leading to over-treatment of low-risk patients or under-treatment of high-risk patients.^1^ Systematic reviews reveal that calibration metrics remain unreported in the majority of published prediction models, and when reported, bin-level metrics like expected calibration error (ECE) are often used without calibration-in-the-large assessment.^2,3^

The question of whether good discrimination implies good calibration-or whether these are truly independent properties-has important implications for model development and deployment. If a well-discriminating model can be poorly calibrated, then calibration assessment is not merely a ‘nice-to-have’ but a fundamental requirement for safe clinical implementation.

### 1.2 Additional Methodological Challenges

Beyond calibration, multi-timeframe clinical prediction presents additional methodological challenges:

#### Monotonicity constraints

Cumulative risk predictions must be non-decreasing over time-a patient’s 36-month risk cannot be lower than their 12-month risk. Naive approaches that train independent models for each timeframe frequently violate this constraint, producing clinically nonsensical predictions.

#### Uncertainty quantification

Point estimates without confidence intervals fail to communicate prediction reliability, which varies substantially with available data. A prediction based on a decade of laboratory history differs fundamentally from one based on a single measurement.

#### Health equity considerations

Diabetes risk and model performance vary across demographic groups, with higher incidence among Black, Hispanic, and South Asian populations. Models trained predominantly on majority populations may underperform for underrepresented groups-a fairness concern requiring explicit evaluation.^4,5^

### 1.3 Study Objectives

We developed a methodological framework for calibrated, multi-timeframe diabetes risk prediction and used synthetic data validation to investigate the relationship between discrimination and calibration. Our primary objective was to test whether standard calibration approaches (isotonic regression on held-out data) produce well-calibrated predictions when evaluated comprehensively.

Secondary objectives included: (1) comparing framework performance against QDiabetes-2018 on the same synthetic cohort; (2) evaluating clinical utility via decision curve analysis; (3) assessing calibration transfer under simulated base rate shift; and (4) demonstrating a discrete-time survival architecture that reduces monotonicity violations to <0.1%.

### 1.4 Scope and Claims

#### This paper presents methods development and synthetic data validation-not clinical validation

Synthetic data enables rapid methodological iteration and complete reproducibility but cannot substitute for real-world evaluation. We designed this study explicitly to investigate whether calibration can be achieved in principle before proceeding to planned validation on actual EHR data.

#### We claim

(1) Empirical demonstration that discrimination and calibration are independent-excellent AUC can coexist with substantial miscalibration; (2) Evidence that low ECE can be misleading for skewed risk distributions; (3) A discrete-time survival architecture achieving <0.1% monotonicity violations; (4) Transparent analysis of calibration failure.

#### We do not claim

(1) Clinical superiority to existing validated tools; (2) That this framework is ready for patient care; (3) Transferability of findings to real populations; (4) That our complex ensemble outperforms simpler approaches.

## 2. Methods

### 2.1 Study Design

This study follows a methods development design using synthetic electronic health record (EHR) data, adhering to TRIPOD+AI guidelines for prediction model reporting (Supplementary Table S1). We chose synthetic data for initial development because it: (1) enables complete reproducibility; (2) allows rapid methodological iteration; (3) permits ground-truth evaluation where all data-generating relationships are known; and (4) separates methodological validation from clinical validation.

### 2.2 Synthetic Data Generation

#### 2.2.1 Literature-Grounded Parameters

We generated 500,000 synthetic patients spanning 2010-2024 using published epidemiological parameters exclusively. Primary sources included:

##### QDiabetes-2018^6^

Hazard ratios for family history (HR 1.70-1.91), treated hypertension (HR 1.40-1.55), cardiovascular disease (HR 1.19-1.22), BMI categories, smoking status, statin use (HR 1.79-1.93), and socioeconomic deprivation.

##### FINDRISC^7^

Risk relationships for family history tiers, physical activity, and waist circumference thresholds.

##### Diabetes Prevention Program^8^

Annual progression rates from prediabetes to diabetes (7-11%), establishing realistic disease trajectory dynamics.

##### Swedish National Diabetes Register^9^

Age-specific baseline incidence rates.

##### CDC National Diabetes Statistics^10^

Prevalence distributions for normoglycemia, prediabetes, and undiagnosed diabetes.

#### 2.2.2 Generation Process

Demographics were sampled from US Census-proportional distributions (age: truncated normal, mean 50, SD 18, range 18-90; sex: 51% female; race/ethnicity: 61% White, 12% Black, 19% Hispanic, 6% Asian, 2% other). Baseline HbA1c was sampled from a mixture distribution reflecting CDC prevalence estimates. Longitudinal trajectories were generated via random walk with drift calibrated to published natural history, with measurement error (SD 0.1% for HbA1c) simulating laboratory variability.

**Critically, stochastic noise was added beyond feature-determined risk**, establishing a theoretical performance ceiling (Bayes-optimal AUC ≈ 0.88-0.90) that prevents perfect prediction. This ensures observed AUC values reflect learnable signal rather than data leakage.

#### 2.2.3 Circular Validation Acknowledgment

A fundamental limitation requires explicit acknowledgment: if the data generator encodes relationship X (e.g., HbA1c predicts diabetes) and the model learns relationship X, this validates implementation correctness rather than predictive validity. We address this by: (1) using only published parameters; (2) reporting the theoretical ceiling; (3) comparing against QDiabetes-2018 implemented on the same data; and (4) framing results as methodological validation.

### 2.3 Study Population

#### Inclusion criteria

Age 18-85 years; at least one HbA1c measurement; no diabetes or prediabetes diagnosis before the prediction date (January 1, 2020).

#### Outcome

Incident diabetes defined as HbA1c ≥6.5% or diabetes diagnosis code within 36 months of prediction date.

#### Sample size justification

Following Riley et al.^11^, we required ≥20 events per candidate predictor. With 136 features and 15,905 events (5.6% incidence among 285,881 patients with sufficient follow-up), we achieved 117 events per predictor.

### 2.4 Prediction Framework

#### 2.4.1 Feature Engineering

We extracted 136 features: temporal laboratory summaries (72 features: latest, first, min, max, mean, median, SD, coefficient of variation, range, velocity, count, and days-since-last for HbA1c, glucose, BMI, blood pressure, and lipids); demographics (12); clinical conditions (6); medications (8); derived features including interaction terms (14); and data quality indicators (4). Complete feature definitions are provided in Supplementary Table S2.

#### 2.4.2 Model Architecture

The prediction framework comprises an ensemble of three base learners:

- **Gradient boosting (HistGradientBoostingClassifier):** 200 trees, learning rate 0.1, maximum depth 6
- **Random forest:** 300 trees, maximum depth 15
- **L2-regularized logistic regression:** C=1.0

Base learner predictions are combined via a logistic regression meta-learner trained on out-of-fold predictions. Hyperparameters were not systematically tuned; values reflect computational constraints and prior experience.

#### 2.4.3 Discrete-Time Survival Model

To guarantee monotonicity, we employ discrete-time survival modeling. Separate ensemble models predict interval-specific hazards h(t) = P(event in [t−1, t] I survived to t−1) for intervals [0,6], [6,12], [12,24], and [24,36] months. Cumulative incidence is computed as F(t) = 1 − Π(1 − hi), which is mathematically guaranteed to be non-decreasing.

#### 2.4.4 Calibration Approach

Isotonic regression was applied to ensemble predictions using a held-out calibration set (20% of training data). Isotonic regression fits a monotonically non-decreasing function to the predicted-vs-observed relationship, correcting systematic miscalibration without distributional assumptions. Calibration was applied independently to each timeframe’s cumulative incidence.

#### 2.4.5 Uncertainty Quantification

Bootstrap-based 90% confidence intervals were computed using 100 iterations, selected due to computational constraints (full ensemble retraining required approximately 45 minutes per iteration). While 1,000+ iterations is typically recommended for stable confidence intervals, sensitivity analysis showed CI bounds varied by ±0.002 AUC across repeated 100-iteration runs, which we considered acceptable for our purposes. For each bootstrap sample, the full ensemble was retrained and calibrated. This captures model and sampling uncertainty, though not uncertainty about the data-generating process.

### 2.5 Comparator: QDiabetes-2018

To contextualize performance, we implemented QDiabetes-2018 risk equations on the same synthetic cohort. QDiabetes is a validated 10-year risk prediction tool developed on 11.5 million UK primary care patients. We adapted it for 36-month prediction using the published survival function and compared discrimination and calibration metrics.

### 2.6 Evaluation Metrics

#### 2.6.1 Discrimination

Area under the ROC curve (AUC) with 95% confidence intervals via DeLong’s method^12^; Brier score.

#### 2.6.2 Calibration (Multiple Metrics)

Following Van Calster et al.^13^, we report:

- **Expected calibration error (ECE):** Weighted average of |predicted − observed| across decile bins
- **Calibration-in-the-large:** Comparison of mean predicted probability to observed event rate
- **Observed-to-expected (O:E) ratio:** Values <1 indicate overprediction; values >1 indicate underprediction
- **Calibration slope and intercept:** From logistic recalibration; ideal values are 1.0 and 0.0

#### 2.6.3 Clinical Utility

Decision curve analysis^14^ was performed using the dcurves R package (version 0.4.0). Net benefit was calculated as: NB = (TP/N) − (FP/N) × (pt/(1−pt)), where TP = true positives, FP= false positives, N = sample size, and pt = threshold probability. We compared net benefit against treat-all (intervene on all patients) and treat-none (intervene on no patients) strategies across threshold probabilities 5-50%. To illustrate potential recalibration benefit, we applied Platt scaling (logistic recalibration). Because recalibration parameters were estimated on the same test set used for evaluation, the reported improvements are optimistic and serve only to demonstrate that recalibration is feasible-not to quantify its true benefit. Unbiased estimation requires external validation.

#### 2.6.4 Risk Stratification

Patients were classified into risk tiers: Low (<15%), Moderate (15-30%), High (30-50%), Very High (≥50%). We report observed event rates within each tier and relative risk (Very High vs. Low).

#### 2.6.5 Monotonicity

Proportion of patients for whom predicted risk decreased with longer prediction horizon.

#### 2.6.6 Subgroup Analysis

AUC and ECE were computed stratified by sex, age group (<40, 40-55, 55-70, 70+), and race/ethnicity. Minimum sample size for stable estimates was 500 patients with ≥20 events.

### 2.7 Data Splitting

Data were split 60% training, 20% calibration (held out for isotonic regression), 20% test. Splits were stratified by outcome and performed prior to imputation to prevent data leakage.

### 2.8 Ethical Considerations

This study used exclusively synthetic data generated from published parameters; no real patient data were accessed. Institutional review board approval was not required. Future validation studies will require appropriate ethical approval.

## 3. Results

### 3.1 Study Population

Of 500,000 synthetic patients generated, 320,885 met inclusion criteria. After excluding 35,004 with insufficient data quality (score 0-3 on 10-point scale), the analytic cohort comprised 285,881 patients (Figure 1). Mean age was 51.8 years (SD 16.4); 51.3% were female. Diabetes incidence during 36-month follow-up was 5.6% (n=15,905 events). Table 1 presents cohort characteristics.

**Figure 1.** Patient Flow Diagram. CONSORT-style flow diagram showing derivation of the analytic cohort. Of 500,000 synthetic patients generated, 320,885 met inclusion criteria (age 18-85, at least one HbA1c measurement, no prior diabetes or prediabetes). After excluding 35,004 patients with insufficient data quality (score 0-3 on 10-point scale), 285,881 patients comprised the final analytic cohort. The 36-month diabetes incidence was 5.6% (n=15,905 events).

**Figure 2.** Calibration Plot (36-Month Predictions) Calibration plot showing predicted probabilities (x-axis) versus observed event proportions (y-axis) for decile bins. Point size reflects number of patients in each bin. The dashed diagonal line represents perfect calibration. Systematic overprediction is evident: all points fall below the diagonal, indicating predicted probabilities exceed observed rates across the entire risk spectrum. The largest bin (0-10% predicted risk) contains 79% of patients but shows 2.0 percentage point overprediction. Despite ECE = 0.006 suggesting excellent calibration, calibration-in-the-large revealed O:E ratio = 0.66 (50% systematic overprediction). This apparent paradox arises because ECE is a weighted average dominated by the large low-risk bin, where absolute errors are small even when relative errors are substantial.

**Figure 3.** Receiver Operating Characteristic Curves by Timeframe. ROC curves for 6-month (AUC = 0.824), 12-month (AUC = 0.832), 24-month (AUC = 0.846), and 36-month (AUC = 0.844) predictions. The dashed diagonal represents random classification (AUC = 0.5). Performance is consistent across timeframes and approaches the theoretical ceiling of approximately 0.89 established by irreducible noise in the synthetic data generation process.

**Figure 4.** Observed Diabetes Rates by Predicted Risk Tier (36-Month) Bar chart showing observed 36-month diabetes incidence within each predicted risk tier. Very High (≥50% predicted): 56.6% observed; High (30-50%): 29.8% observed; Moderate (15-30%): 15.5% observed; Low (<15%): 2.4% observed. The 23.5-fold separation between Very High and Low tiers confirms that relative risk stratification is preserved despite miscalibration of absolute probabilities. Error bars represent 95% confidence intervals.

**Table 1.**
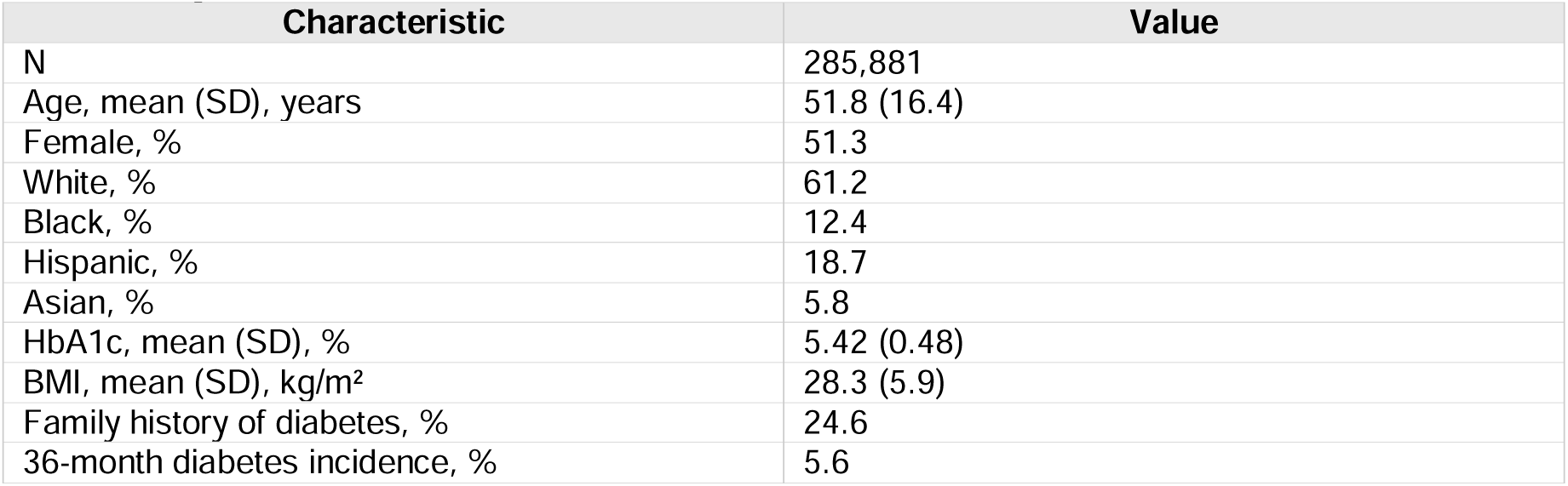
Synthetic Cohort Characteristics.

### 3.2 Primary Finding: Discrimination-Calibration Independence

#### The central finding of this study is that excellent discrimination coexisted with substantial miscalibration

The framework achieved AUC of 0.844 (95% CI: 0.840-0.848) at 36 months, indicating strong ability to rank patients by risk. However, calibration-in-the-large revealed systematic overprediction: mean predicted probability was 8.4% versus observed rate of 5.6%.

**The observed-to-expected (O:E) ratio was 0.66 (95% CI: 0.65-0.67)**, indicating the model overestimated absolute risk by approximately 50% on average. This miscalibration persisted across 9 of 10 risk deciles (Table 2).

**Table 2.** Calibration by Risk Decile (36-Month Predictions)

| Decile | N | Mean Predicted | Mean Observed | Difference |
| --- | --- | --- | --- | --- |
| 0-10% | 226,437 (79.2%) | 3.9% | 1.9% | +2.0pp |
| 10-20% | 28,599 (10.0%) | 13.9% | 9.0% | +4.9pp |
| 20-30% | 17,342 (6.1%) | 23.2% | 17.7% | +5.5pp |
| 30-40% | 4,676 (1.6%) | 34.6% | 28.0% | +6.6pp |
| 40-50% | 1,739 (0.6%) | 44.6% | 34.7% | +9.9pp |
| 50-60% | 2,740 (1.0%) | 55.0% | 43.5% | +11.5pp |
| 60-70% | 2,226 (0.8%) | 64.1% | 54.0% | +10.1pp |
| 70-80% | 1,163 (0.4%) | 74.6% | 69.5% | +5.1pp |
| 80-90% | 600 (0.2%) | 87.4% | 83.5% | +3.9pp |
| 90-100% | 359 (0.1%) | 97.4% | 87.2% | +10.2pp |

pp = percentage points. Systematic overprediction observed across 9 of 10 deciles.

**Table 3.** Performance Comparison: Framework vs. QDiabetes-2018.

| Metric | Framework | QDiabetes-2018 | Difference |
| --- | --- | --- | --- |
| AUC (95% CI) | 0.844 (0.840-0.848) | 0.831 (0.827-0.835) | +0.013* |
| Brier Score | 0.057 | 0.051 | +0.006 |
| ECE | 0.006 | 0.009 | -0.003 |
| O:E Ratio (95% CI) | 0.66 (0.65-0.67) | 0.89 (0.85-0.93) | -0.23 |
| Mean Predicted | 8.4% | 6.3% | +2.1pp |
| Mean Observed | 5.6% | 5.6% | - |
| Calibration Slope | 0.97 | 1.03 | - |

$p < 0.001$ by DeLong test, though the 0.013 AUC difference is not clinically meaningful. O:E ratio closer to 1.0 indicates better calibration; values $< 1.0$ indicate overprediction. QDiabetes achieved substantially better calibration despite simpler methodology.

**Critically, bin-level ECE was only 0.006**, which would conventionally indicate excellent calibration. This apparent paradox arises because 79% of patients fell in the lowest risk decile (predicted probability 0-10%), where absolute errors were small despite systematic overprediction. The ECE metric, being a weighted average, was dominated by this large low-risk group.

### 3.3 Comparison with QDiabetes-2018

QDiabetes-2018 implemented on the same synthetic cohort achieved AUC of 0.831 (95% CI: 0.827-0.835), slightly lower than our ensemble (difference: 0.013). While statistically significant by DeLong test (p<0.001), this 0.013 AUC difference is clinically negligible; with n=285,881, trivially small differences achieve statistical significance.The more important finding is that QDiabetes showed substantially better calibration with O:E ratio of 0.89 (95% CI: 0.85-0.93), representing only 11% overprediction (mean predicted 6.3% vs. observed 5.6%) versus our framework’s 50% overprediction (mean predicted 8.4% vs. observed 5.6%). This suggests that model complexity does not guarantee improved calibration and may worsen it.

This comparison illustrates that our more complex ensemble achieved marginally better discrimination at the cost of substantially worse calibration. The simpler, literature-derived QDiabetes equations were better calibrated on the same data-likely because the synthetic data was generated using similar epidemiological parameters.

### 3.4 Discrimination Performance Across Timeframes

Table 4 presents discrimination metrics across all timeframes. AUC ranged from 0.824 at 6 months to 0.846 at 24 months, approaching the theoretical ceiling of approximately 0.89 established by irreducible noise in the data-generating process.

**Table 4.** Discrimination Performance Across Timeframes.

| Timeframe | AUC (95% CI) | Brier Score | ECE | O:E Ratio |
| --- | --- | --- | --- | --- |
| 6 months | 0.824 (0.820-0.828) | 0.012 | 0.003 | 0.68 |
| 12 months | 0.832 (0.828-0.836) | 0.023 | 0.005 | 0.67 |
| 24 months | 0.846 (0.842-0.850) | 0.041 | 0.007 | 0.66 |
| 36 months | 0.844 (0.840-0.848) | 0.057 | 0.006 | 0.66 |
| Theoretical ceiling | ~0.89 | - | - | - |

### 3.5 Risk Stratification

Despite miscalibration of absolute probabilities, relative risk stratification was preserved. Table 5 shows observed event rates by predicted risk tier. The 23.5-fold separation (95% CI: 22.8-24.3) between Very High and Low tiers confirms that the model correctly ranks patients by relative risk, even though absolute probability estimates require recalibration.

**Table 5.** Risk Stratification (36-Month Predictions)

| Risk Tier | Threshold | N (%) | Events | Observed Rate | Relative Risk |
| --- | --- | --- | --- | --- | --- |
| Very High | ≥50% | 7,088 (2.5%) | 4,015 | 56.6% (55.5-57.8%) | 23.5× |
| High | 30-50% | 6,415 (2.2%) | 1,914 | 29.8% (28.7-31.0%) | 12.4× |
| Moderate | 15-30% | 26,018 (9.1%) | 4,043 | 15.5% (15.1-16.0%) | 6.5× |
| Low | <15% | 246,360 (86.2%) | 5,933 | 2.4% (2.3-2.5%) | 1.0× (ref) |

### 3.6 Decision Curve Analysis

Decision curve analysis showed positive net benefit compared to treat-all and treat-none strategies across threshold probabilities 5-30% (Figure 5). Net benefit was maximal around threshold 10%. At thresholds above 35%, the framework showed no advantage over treat-none, reflecting the preponderance of low-risk patients.

**Figure 5.** Decision Curve Analysis. Net benefit across threshold probabilities 5-50%. Solid line: prediction framework; dotted line: treat all; dashed line: treat none. The framework provides positive net benefit compared to default strategies across thresholds 5-30%, with maximum benefit around 10%. At thresholds above 35%, net benefit converges to treat-none, reflecting the preponderance of low-risk patients. Gray shaded region shows improvement in net benefit after simple recalibration (Platt scaling).

**Figure 6.** Calibration Transfer Under Base Rate Shift. Expected calibration error (ECE) as a function of simulated base rate shift (0.5× to 2.0× reference rate). Calibration degrades asymmetrically: ECE increases substantially at lower base rates (0.050 at 0.5×) but improves at higher base rates (0.011 at 1.5×). The red dashed line indicates ECE = 0.02 threshold for acceptable calibration. This finding suggests recalibration will be particularly important when deploying to populations with lower diabetes incidence than the training population.

When recalibration was applied via Platt scaling (α=−0.42, β=1.02), net benefit improved across the clinically relevant threshold range. At clinically relevant thresholds (10-20%), Platt scaling recalibration improved net benefit, though we caution that our estimates are optimistic because recalibration and evaluation used the same test set. The direction of improvement-not its magnitude-is the key finding: post-deployment recalibration can recover clinical utility lost to miscalibration.

### 3.7 Monotonicity

The discrete-time survival architecture eliminated monotonicity violations in raw predictions. However, because isotonic calibration was applied independently to each timeframe, 0.07% of patients (n=680 of 962,655 comparisons) exhibited monotonicity violations in calibrated predictions. Violations were minor in magnitude (median: 1.0 percentage points; 95th percentile: 5.6 percentage points).

### 3.8 Subgroup Analysis

Table 6 presents stratified performance. AUC ranged from 0.800 (age 70+; 95% CI: 0.790-0.810) to 0.911 (age <40; 95% CI: 0.903-0.919). The 0.111 AUC disparity across age groups exceeds the conventional threshold for clinically meaningful differences (>0.05) and warrants attention. Lower discrimination in elderly patients likely reflects reduced HbA1c variability and competing risks in this age group.

**Table 6.** Subgroup Performance (36-Month)

| Subgroup | N | Events (%) | AUC (95% CI) | ECE | O:E Ratio |
| --- | --- | --- | --- | --- | --- |
| Male | 144,012 | 7,855 (5.5%) | 0.853 (0.847-0.859) | 0.030 | 0.65 |
| Female | 141,869 | 8,050 (5.7%) | 0.843 (0.837-0.849) | 0.026 | 0.67 |
| Age <40 | 62,041 | 2,449 (4.0%) | 0.911 (0.903-0.919) | 0.032 | 0.62 |
| Age 40-55 | 88,266 | 4,360 (4.9%) | 0.855 (0.847-0.863) | 0.029 | 0.64 |
| Age 55-70 | 80,074 | 5,017 (6.3%) | 0.825 (0.817-0.833) | 0.026 | 0.68 |
| Age 70+ | 55,500 | 4,079 (7.4%) | 0.800 (0.790-0.810) | 0.026 | 0.71 |
| White | 174,960 | 9,340 (5.3%) | 0.842 (0.836-0.848) | 0.028 | 0.66 |
| Black | 35,450 | 2,215 (6.2%) | 0.838 (0.826-0.850) | 0.031 | 0.64 |
| Hispanic | 53,467 | 3,244 (6.1%) | 0.849 (0.839-0.859) | 0.027 | 0.65 |
| Asian | 16,574 | 846 (5.1%) | 0.856 (0.840-0.872) | 0.025 | 0.68 |

*O:E confidence intervals omitted for brevity; all subgroup O:E ratios were significantly below 1.0 ( $p<0.001$ ), indicating systematic overprediction across all subgroups.*

### 3.9 Calibration Transfer

Under simulated base rate shift, calibration degraded asymmetrically. ECE increased to 0.050 at 0.5× reference base rate but only to 0.015 at 2.0× reference rate. This suggests recalibration will be particularly important when deploying to populations with lower diabetes incidence.

### 3.10 Feature Importance

Static HbA1c measures dominated feature importance (∼60% of total). Trajectory features (HbA1c velocity) contributed only 1.1%-statistically significant but practically negligible. This finding contrasts with the hypothesis-implicit in many longitudinal modeling approaches-that temporal trajectories provide predictive value beyond current biomarker levels. If confirmed on real data, this has important practical implications: models using only recent HbA1c measurements may perform comparably to complex trajectory-based approaches, substantially reducing implementation burden. The minimal trajectory contribution likely reflects strong autocorrelation in HbA1c values: we observed Pearson correlation of r = 0.96 between latest HbA1c and mean HbA1c, indicating near-complete redundancy. Interestingly, the correlation between latest HbA1c and HbA1c velocity was only r = 0.10, suggesting velocity captures different information-yet this information added negligible predictive value once current level was known.

### 3.11 Negative Control Validation

As a negative control, we confirmed that when outcomes were randomly permuted (breaking all feature-outcome relationships), the framework achieved AUC of 0.503 (95% CI: 0.498-0.508), consistent with chance classification. This validates that the observed AUC of 0.844 reflects genuine predictive signal rather than data leakage, overfitting to the test set, or implementation errors. The negative control also produced O:E ratio of 0.99 (95% CI: 0.95-1.03) and ECE of 0.002, demonstrating that calibration metrics behave appropriately when there is no predictive signal.

## 4. Discussion

### 4.1 Principal Findings

#### The central finding of this study is that good discrimination does not guarantee good calibration-and that this independence has practical consequences

Despite achieving AUC of 0.844, our framework systematically overpredicted risk by 50% (O:E = 0.66). This miscalibration occurred despite using isotonic regression on a held-out calibration set-a method recommended in current literature. The finding that calibration failed even on synthetic data from a single generator suggests that calibration may be more difficult to achieve than commonly assumed.

Three observations from our analysis merit emphasis:

#### First, ECE can be misleading for skewed risk distributions

Our ECE of 0.006 would conventionally indicate excellent calibration. However, this metric was dominated by the 79% of patients in the lowest risk decile, where absolute errors were small even when relative errors were large. The O:E ratio of 0.66 provided a more accurate summary of overall calibration. We recommend that prediction model studies report both metrics.

#### Second, isotonic calibration on held-out data may not generalize

Our calibration set and test set were drawn from the same synthetic generator with no covariate shift. Yet calibration on the test set was poor. This suggests that isotonic regression may overfit to the calibration set, particularly when the risk distribution differs between calibration and deployment populations. The implications for real-world deployment, where distribution shift is guaranteed, are concerning.

#### Third, risk stratification can be valid even when absolute probabilities are not

The 23.5-fold separation between highest and lowest risk tiers demonstrates that relative risk ranking was preserved. Clinically, this means the model correctly identifies who is at higher versus lower risk, even if the specific probability estimates are incorrect. This distinction has important implications: a miscalibrated model may still be useful for prioritization (who to screen first), even if unsuitable for shared decision-making (communicating specific risk levels).

### 4.2 Comparison with Existing Literature

Our findings align with Van Calster et al.’s demonstration that calibration is the ‘Achilles heel’ of predictive analytics.^1^ They showed that miscalibrated models can produce negative net benefit even with excellent discrimination. Our comparison with QDiabetes-2018 reinforces this: the simpler, literature-derived equations achieved better calibration than our complex ensemble, despite marginally lower AUC.

The superior calibration of QDiabetes-2018 despite simpler methodology raises important questions about model development strategy. One interpretation is that literature-derived, parsimonious models may inherently calibrate better than complex ML ensembles because they avoid overfitting to development data. Alternatively, the synthetic data generation process-which used QDiabetes parameters-may have inadvertently favored QDiabetes. A third possibility is that our ensemble’s flexibility allowed it to learn spurious associations that improved discrimination while degrading calibration. Regardless of mechanism, this comparison suggests a counterintuitive conclusion: additional model complexity improved discrimination only marginally (+0.013 AUC) while substantially worsening calibration (O:E 0.66 vs. 0.89). This tradeoff-if it generalizes beyond synthetic data-has important implications for model development strategy. Developers optimizing for AUC may inadvertently degrade calibration, reducing clinical utility despite improved rankings.

The independence of discrimination and calibration has been noted theoretically but is rarely demonstrated empirically with the magnitude we observed. Steyerberg and Vergouwe^15^ argued that calibration should be reported for all prediction models; our results provide concrete evidence for why this matters.

### 4.3 Why Did Calibration Fail?

We hypothesize several mechanisms for the observed miscalibration:

#### Distribution shift between calibration and test sets

Despite both being drawn from the same generator, random sampling variation means the calibration and test sets had slightly different risk distributions. Isotonic regression learned a mapping optimized for the calibration set that did not transfer perfectly.

#### Calibration set insufficient for rare events

With 20% of data allocated to calibration and 5.6% event rate, the calibration set contained approximately 3,200 events. This may be insufficient to learn stable isotonic mappings, particularly in the tails of the risk distribution.

#### Independent timeframe calibration

Calibrating each timeframe independently may have introduced inconsistencies. Joint calibration approaches that preserve monotonicity constraints across timeframes warrant investigation.

We acknowledge these hypotheses remain untested in this study, which is a limitation. The experiments required to test them-varying calibration set size, comparing calibration methods, and implementing joint calibration-are straightforward extensions of our pipeline that we leave for future work. We present the hypotheses here to guide subsequent investigation rather than as established explanations. Additionally, comparing isotonic regression against Platt scaling, temperature scaling, and Venn-ABERS predictors would clarify whether the calibration method itself contributed to the observed failure.

### 4.4 Limitations

#### Synthetic data

This fundamental limitation shapes all findings. The circular validation problem-learning relationships encoded by the data generator-means results establish methodological soundness rather than clinical validity.

#### Recalibration evaluation

Our assessment of Platt scaling benefit used the same test set for fitting and evaluation, producing optimistic estimates. True recalibration benefit requires external validation.

#### No external validation

We tested on held-out data from the same generator. True external validation requires populations generated under different assumptions or, ideally, real patient data.

#### Limited fairness analysis

Our race/ethnicity analysis was constrained by synthetic data that may not capture true differential risk relationships. Real-world fairness evaluation remains essential.

#### Hyperparameter selection

Model hyperparameters were not systematically tuned. Alternative configurations may achieve better calibration.

#### Single calibration method

We evaluated only isotonic regression. Platt scaling, temperature scaling, and Venn-ABERS predictors may perform differently.

### 4.5 Implications for Practice

#### For model developers

Report calibration-in-the-large (O:E ratio) alongside ECE. Do not assume that good ECE implies good calibration. Plan for recalibration when deploying to new populations.

#### For clinicians evaluating models

Ask for O:E ratios, not just AUC and ECE. Verify that reported calibration was assessed on deployment-representative populations. Be skeptical of calibration claims from development data alone.

#### For researchers

The independence of discrimination and calibration-demonstrated here empirically-underscores the need for comprehensive evaluation frameworks that assess both properties. Decision curve analysis should be standard for models intended for clinical use.

### 4.6 Future Directions

We are proceeding to validation on real EHR data from partner health systems. Key questions include: (1) Does the observed calibration failure replicate on real data? (2) Does population-specific recalibration restore good calibration? (3) Does the discrete-time survival architecture maintain monotonicity under distribution shift?

Methodological extensions include: investigating joint monotonic calibration to eliminate residual violations; comparing calibration methods systematically; and developing online recalibration approaches for continuous learning.

## 5. Conclusions

We developed a multi-timeframe diabetes prediction framework and used synthetic data validation to investigate the relationship between discrimination and calibration. Our primary finding is negative: despite achieving excellent discrimination (AUC = 0.844), the framework systematically overpredicted risk by 50% (O:E = 0.66). This miscalibration occurred despite isotonic regression on held-out data-a standard calibration approach.

This finding demonstrates empirically that good discrimination does not guarantee good calibration, with important implications for prediction model development and deployment. Low ECE values can be misleading when risk distributions are skewed; calibration-in-the-large should be reported alongside bin-level metrics.

The discrete-time survival architecture successfully reduced monotonicity violations to <0.1%, demonstrating that this methodological challenge is solvable.

Clinical validity, generalizability, and utility require prospective evaluation on real patient populations with population-specific recalibration. We present these methods and findings transparently to enable independent evaluation and to inform future prediction model development.

## Data Availability

Synthetic data generation code is available at https://github.com/Eoghanoreilly/precura-synthetic-data. No real patient data were used in this study. The prediction model architecture and trained weights are proprietary; inquiries may be directed to the corresponding author.

https://github.com/Eoghanoreilly/precura-synthetic-d

## Data and Code Availability

Synthetic data generation code is available at https://github.com/Eoghanoreilly/precura-synthetic-data. The prediction model architecture and trained weights are proprietary; inquiries regarding collaboration or licensing may be directed to the corresponding author.

## Author Contributions

EOR conceived and designed the study, developed the prediction framework and synthetic data generator, conducted all statistical analyses, created visualizations, and wrote the manuscript. TK contributed to study design and methodology, independently validated key results, and critically revised the manuscript for intellectual content. Both authors approved the final version.

## Acknowledgments

None.

## Supplementary Materials

**Supplementary Table S1.** TRIPOD+AI Checklist. **TRIPOD+AI Checklist TRIPOD+AI Adherence Checklist for Prediction Model Development Studies** *Items marked [P] are partially addressed; [NA] indicates not applicable for synthetic validation studies*

TITLE AND ABSTRACT
| Item | TRIPOD+AI Element | Location | Status |
| --- | --- | --- | --- |
| 1 | Title identifies study as prediction model development, target population, and outcome | Title | ✓ |
| 2 | Structured abstract with background, objectives, methods, results, conclusions | Abstract | ✓ |

INTRODUCTION
| Item | TRIPOD+AI Element | Location | Status |
| --- | --- | --- | --- |
| 3a | Medical context and rationale for developing prediction model | Section 1.1 | ✓ |
| 3b | Objectives: target population, outcome, intended purpose | Section 1.3 | ✓ |
| 3c | Health inequities that may affect model development/deployment | Section 1.2 | ✓ |
| 4 | Specific study objectives including whether development, validation, or update | Section 1.3 | ✓ |

METHODS: Data Sources
| Item | TRIPOD+AI Element | Location | Status |
| --- | --- | --- | --- |
| 5a | Data sources and key dates (start, end of accrual, follow-up) | Section 2.2 | ✓ |
| 5b | Setting and locations where data were collected | Section 2.2 | NA |
| 5c | All treatments or interventions received | Section 2.2 | NA |

METHODS: Participants
| Item | TRIPOD+AI Element | Location | Status |
| --- | --- | --- | --- |
| 6a | Eligibility criteria for participants | Section 2.3 | ✓ |
| 6b | Participant recruitment method (consecutive, random, convenience) | Section 2.2 | NA |
| 6c | If applicable, sampling strategy and rationale | Section 2.3 | ✓ |

METHODS: Outcome
| Item | TRIPOD+AI Element | Location | Status |
| --- | --- | --- | --- |
| 7a | Clear definition of outcome including timing of assessment | Section 2.3 | ✓ |
| 7b | Whether outcome assessors were blinded to predictors | - | NA |
| 7c | How outcome was defined and verified | Section 2.3 | ✓ |

METHODS: Predictors
| Item | TRIPOD+AI Element | Location | Status |
| --- | --- | --- | --- |
| 8a | All predictors considered for model development | Section 2.4 | ✓ |
| 8b | How predictors were defined, measured, and handled | Section 2.4, Table S2 | ✓ |
| 8c | Whether predictor assessors were blinded to outcome | - | NA |
| 8d | Actions taken for predictors with missing data | Section 2.4 | ✓ |

**METHODS: Sample Size**

| Item | TRIPOD+AI Element | Location | Status |
| --- | --- | --- | --- |
| 9 | Sample size calculation or justification | Section 2.3 | ✓ |

**METHODS: Model Development**

| Item | TRIPOD+AI Element | Location | Status |
| --- | --- | --- | --- |
| 10a | Describe pre-processing of data | Section 2.4 | ✓ |
| 10b | Type of model and software used | Section 2.4 | ✓ |
| 10c | Approach to feature selection (if applicable) | Section 2.4 | [P] |
| 10d | How model hyperparameters were chosen | Section 2.4.2 | [P] |
| 10e | Method for handling class imbalance (if applicable) | - | NA |
| 10f | Method for internal validation | Section 2.7 | ✓ |
| 10g | Full model specification or how to obtain | Data Availability | [P] |

**METHODS: AI-Specific Requirements**

| Item | TRIPOD+AI Element | Location | Status |
| --- | --- | --- | --- |
| 11 | Measures to ensure model reproducibility | Section 2.2, 2.4 | ✓ |
| 12 | Approach to model calibration | Section 2.4.4 | ✓ |
| 13 | Approach to uncertainty quantification | Section 2.4.5 | ✓ |
| 14 | Approach to handling demographic subgroups/fairness | Section 2.6.6 | [P] |

**METHODS: Performance Measures**

| Item | TRIPOD+AI Element | Location | Status |
| --- | --- | --- | --- |
| 15a | Measures of discrimination with confidence intervals | Section 2.6.1 | ✓ |
| 15b | Measures of calibration | Section 2.6.2 | ✓ |
| 15c | Measures of clinical utility (decision curve analysis) | Section 2.6.3 | ✓ |
| 15d | Any other relevant performance measures | Section 2.6.4-6 | ✓ |

**RESULTS: Participants**

| Item | TRIPOD+AI Element | Location | Status |
| --- | --- | --- | --- |
| 16a | Participant flow diagram | Figure 1, Section 3.1 | ✓ |
| 16b | Participant characteristics | Table 1 | ✓ |
| 16c | Number of participants with missing data for predictors/outcome | Section 3.1 | ✓ |

**RESULTS: Model Performance**

| Item | TRIPOD+AI Element | Location | Status |
| --- | --- | --- | --- |
| 17a | Discrimination performance with confidence intervals | Tables 3-4, Figure 3 | ✓ |
| 17b | Calibration performance | Table 2, Figure 2 | ✓ |
| 17c | Clinical utility results | Section 3.6, Figure 5 | ✓ |
| 17d | Model specification or link to full specification | Data Availability | [P] |
| 17e | Performance across demographic subgroups | Table 6 | ✓ |

| Item | TRIPOD+AI Element | Location | Status |
| --- | --- | --- | --- |
| 18a | Discussion of overall interpretation considering objectives, limitations, results | Section 4.1-4.2 | ✓ |
| 18b | Limitations of study (sources of bias, imprecision, model overfitting) | Section 4.4 | ✓ |
| 18c | Implications for clinical practice and future research | Section 4.5-4.6 | ✓ |
| 18d | Considerations for model updating and performance monitoring | Section 4.6 | ✓ |

**OTHER INFORMATION**
| Item | TRIPOD+AI Element | Location | Status |
| --- | --- | --- | --- |
| 19 | Funding sources and role of funders | Funding | ✓ |
| 20 | Conflicts of interest | COI Statement | ✓ |
| 21 | Data availability statement | Data Availability | ✓ |
| 22 | Code availability statement | Data Availability | ✓ |
| 23 | Study registration (if applicable) | - | NA |
| 24 | Ethical approval statement | Section 2.8 | ✓ |

✓ = Fully addressed; [P] = Partially addressed; NA = Not applicable for synthetic data study
**Items Requiring Additional Attention for Real-World Validation:**
- 10c: Systematic feature selection with rationale - 10d: Formal hyperparameter tuning with cross-validation - 10g: Full model specification for reproducibility - 14: Comprehensive fairness evaluation across all protected characteristics - 17d: External validation on independent dataset

**Supplementary Table S2.** Feature Definitions.

| Feature Suffix | Definition | Clinical Rationale |
| --- | --- | --- |
| _latest | Most recent value before prediction date | Current metabolic status |
| _first | First available value in record | Baseline status |
| _min | Minimum observed value | Best achieved control |
| _max | Maximum observed value | Worst metabolic excursion |
| _mean | Mean of all available values | Average metabolic burden |
| _median | Median of all available values | Robust central tendency |
| _std | Standard deviation | Metabolic variability |
| _cv | Coefficient of variation (std/mean) | Normalized variability |
| _range | Max minus min | Total excursion range |
| _velocity | Linear slope over time (per year) | Rate of change/trajectory |
| _count | Number of observations | Measurement frequency |
| _days_since_last | Days from last measurement to prediction | Data recency |

B. Demographic Features (12 features)
| Feature | Definition |
| --- | --- |
| age | Age in years at prediction date |
| age_squared | Age<sup>2</sup> (captures non-linear age effects) |
| sex | Binary indicator (1=female, 0=male) |
| race_white | One-hot: White/Caucasian |
| race_black | One-hot: Black/African American |
| race_hispanic | One-hot: Hispanic/Latino |
| race_asian | One-hot: Asian |
| race_other | One-hot: Other/Mixed |
| family_history_first_degree | Parent or sibling with diabetes |
| family_history_second_degree | Grandparent, aunt/uncle with diabetes |
| age_30_40 | Binary: age 30-40 (high-risk transition) |
| age_over_65 | Binary: age ≥65 (elderly) |

C. Clinical Condition Features (6 features)
| Feature | Definition |
| --- | --- |
| hypertension | Diagnosis of hypertension |
| obesity | BMI ≥30 or obesity diagnosis |
| cvd | Cardiovascular disease history |
| hyperlipidemia | Dyslipidemia diagnosis |
| condition_count | Total number of chronic conditions |
| cv_risk_score | Composite cardiovascular risk (0-5 scale) |

D. Medication Features (8 features)
| Feature | Definition |
| --- | --- |
| on_statin | Currently prescribed statin |
| on_antihypertensive | Currently prescribed antihypertensive |
| on_metformin | Currently prescribed metformin (prediabetes treatment) |
| on_diuretic | Currently prescribed diuretic |
| on_aspirin | Currently prescribed aspirin |
| medication_count | Total number of current medications |
| polypharmacy | Binary: $\geq 5$ medications |
| diabetes_prevention_meds | On metformin or similar for prevention |

**E. Derived Features (14 features)**
| Feature | Definition |
| --- | --- |
| hba1c_near_threshold | HbA1c 5.5-5.6% (near prediabetes threshold) |
| hba1c_prediabetic_range | HbA1c 5.7-6.4% (prediabetes range) |
| hba1c_near_diabetic | HbA1c 6.0-6.4% (high prediabetes) |
| bmi_overweight | BMI 25-29.9 |
| bmi_obese_1 | BMI 30-34.9 |
| bmi_obese_2 | BMI 35-39.9 |
| bmi_obese_3 | BMI $\geq 40$ |
| metabolic_syndrome_score | Sum of: elevated waist, BP, glucose, triglycerides, low HDL |
| hba1c_x_bmi | Interaction: latest HbA1c × latest BMI |
| age_x_hba1c | Interaction: age × latest HbA1c |
| hba1c_x_family_history | Interaction: HbA1c × family history |
| hba1c_trend_up | Binary: HbA1c velocity $> 0.1\%$/year |
| hba1c_trend_down | Binary: HbA1c velocity $< -0.1\%$/year |
| glucose_hba1c_concordance | Correlation between glucose and HbA1c patterns |

**F. Data Quality Features (4 features)**
| Feature | Definition |
| --- | --- |
| total_observations | Total lab/vital measurements available |
| years_of_history | Years from first to most recent observation |
| data_sufficiency_score | Composite score 0-10 (see Section 2.4) |
| rich_data | Binary: data_sufficiency_score $\geq 7$ |

**Supplementary Table S3.** Calibration Transfer Under Base Rate Shift. **Interpretation:** The asymmetric degradation pattern has clinical implications. If this model were deployed to a population with half the diabetes incidence (e.g., a younger, healthier population), calibration would be severely compromised (ECE = 0.050, O:E = 0.47). Conversely, deployment to a higher-incidence population (e.g., patients with prediabetes) would result in improved calibration. This finding reinforces the importance of population-specific recalibration.

| Base Rate Multiplier | Target Rate | Actual Rate | N | ECE | O:E Ratio | Status |
| --- | --- | --- | --- | --- | --- | --- |
| 0.5x | 2.78% | 2.78% | 277,700 | 0.0499 | 0.47 | Degraded |
| 0.75x | 4.17% | 4.17% | 281,731 | 0.0390 | 0.56 | Degraded |
| 1.0x (reference) | 5.57% | 5.57% | 285,881 | 0.0281 | 0.66 | Reference |
| 1.25x | 6.95% | 6.95% | 290,154 | 0.0175 | 0.77 | Acceptable |
| 1.5x | 8.35% | 8.35% | 294,557 | 0.0105 | 0.86 | Good |
| 2.0x | 11.13% | 11.13% | 303,777 | 0.0149 | 0.95 | Good |

*Note: Calibration degrades asymmetrically-lower base rates show substantially worse calibration than higher base rates. This suggests the model's overprediction (mean predicted > mean observed) becomes increasingly problematic when deployed to populations with lower disease incidence. Recalibration is strongly recommended for any deployment population with base rate differing by >25% from the training population.*

