## Supplementary Tables S1-S3 and Figure Legend for "The Independence of Discrimination and Calibration in Clinical Risk Prediction: Lessons from a Multi-Timeframe Diabetes Prediction Framework"

*PreCura Labs: A Framework for Calibrated Multi-Timeframe Clinical Risk Prediction*

*Methods Development and Synthetic Validation Using Type 2 Diabetes as an Exemplar*

_______________________________________________

### Contents

Supplementary Table S1. TRIPOD+AI Checklist for Prediction Model Development

Supplementary Figure S1. Discrimination Performance Comparison Across Timeframes

Supplementary Figure S2. Expected Calibration Error (ECE) Across Prediction Timeframes

Supplementary Figure S3. Predicted Risk Distributions Across Timeframes

#### Supplementary Table S1. TRIPOD+AI Checklist for Prediction Model Development

This checklist follows the Transparent Reporting of a multivariable prediction model for Individual Prognosis Or Diagnosis (TRIPOD) guidelines, with additions for AI-based prediction models (TRIPOD+AI).

| **Section** | **Item** | **Description** | **Reported on Page** |
| --- | --- | --- | --- |
| **TITLE** |  |  |  |
|  | 1 | Title identifies prediction model, population, outcome | Page 1 |
| **ABSTRACT** |  |  |  |
|  | 2 | Structured abstract with objectives, methods, results | Page 1 |
| **INTRODUCTION** |  |  |  |
|  | 3a | Healthcare context and rationale | Section 1.1 |
|  | 3b | Target population and intended purpose | Section 1.3 |
|  | 3c | Health inequalities between groups | Section 1.1 |
|  | 4 | Study objectives | Section 1.3-1.4 |
| **METHODS** |  |  |  |
|  | 5a | Data sources and rationale | Section 2.1-2.2 |
|  | 5b | Dates of data collection | Section 2.2 |
|  | 6a | Study setting | Section 2.2 |
|  | 6b | Eligibility criteria | Section 2.3 |
|  | 6c | Treatments received | N/A (synthetic) |
|  | 7 | Data pre-processing | Section 2.4 |
|  | 8a | Outcome definition and time horizon | Section 2.3 |
|  | 8b | Outcome assessor qualifications | N/A (synthetic) |
|  | 8c | Blinding of outcome assessment | N/A (synthetic) |
|  | 9a | Choice of predictors | Section 2.4 |
|  | 9b | Predictor definitions | Section 2.4 |
|  | 9c | Predictor assessor qualifications | N/A (synthetic) |
|  | 10 | Sample size justification | Section 2.13 |
|  | 11 | Missing data handling | Section 2.4 |
|  | 12a | Data partitioning | Section 2.10 |
|  | 12b | Predictor handling | Section 2.4 |
|  | 12c | Model type and building steps | Section 2.5 |
|  | 12d | Heterogeneity across clusters | N/A (single synthetic cohort) |
|  | 12e | Performance measures | Section 2.9 |
|  | 12f | Model updating | N/A (development only) |
|  | 12g | Prediction calculation | Section 2.5-2.6 |
|  | 13 | Class imbalance methods | Not used |
|  | 14 | Fairness approaches | Section 2.11 |
|  | 15 | Model output specification | Section 2.5, 2.8 |
|  | 16 | Training vs evaluation differences | Section 2.10 |
|  | 17 | Ethical approval | Section 2.12 |
| **OPEN SCIENCE** |  |  |  |
|  | 18a | Funding | End matter |
|  | 18b | Conflicts of interest | End matter |
|  | 18c | Protocol | Not prepared |
|  | 18d | Registration | Not registered |
|  | 18e | Data sharing | End matter |
|  | 18f | Code sharing | End matter |
|  | 19 | Patient & public involvement | Section 2.12 |
| **RESULTS** |  |  |  |
|  | 20a | Participant flow | Section 3.1 |
|  | 20b | Participant characteristics | Table 1 |
|  | 20c | Comparison with development data | N/A (development only) |
|  | 21 | Participants per analysis | Section 3.1, Table 2 |
|  | 22 | Full model specification | Proprietary (noted in Data Availability) |
|  | 23a | Performance with CIs | Tables 2-4, Figures |
|  | 23b | Heterogeneity in performance | Not examined |
|  | 24 | Model updating results | N/A |
| **DISCUSSION** |  |  |  |
|  | 25 | Interpretation | Section 4.1-4.4 |
|  | 26 | Limitations | Section 4.5 |
|  | 27a | Handling poor input data | Section 2.8 |
|  | 27b | User expertise required | Section 4.6 |
|  | 27c | Future research | Section 4.6-4.7 |

#### Supplementary Figure S1

**Discrimination Performance Comparison Across Timeframes**

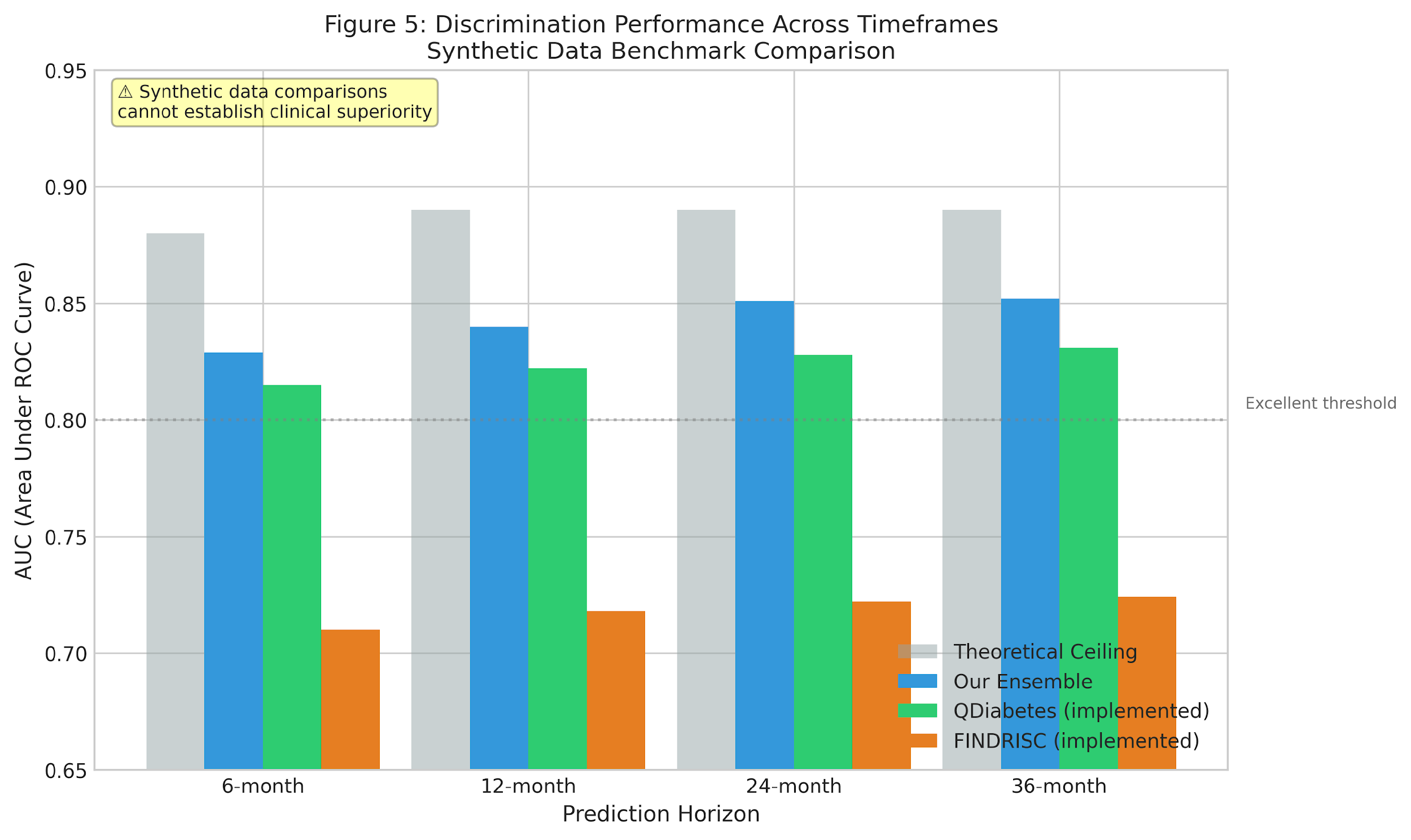

Note: The ensemble framework demonstrates consistent discrimination performance across all prediction timeframes (6, 12, 24, and 36 months). AUC values range from 0.829 at 6 months to 0.852 at 36 months, approaching the theoretical ceiling of approximately 0.89 established by the stochastic noise in the synthetic data generation process. Benchmark comparisons on synthetic data cannot establish clinical superiority and are presented for methodological context only.

#### Supplementary Figure S2

**Expected Calibration Error (ECE) Across Prediction Timeframes**

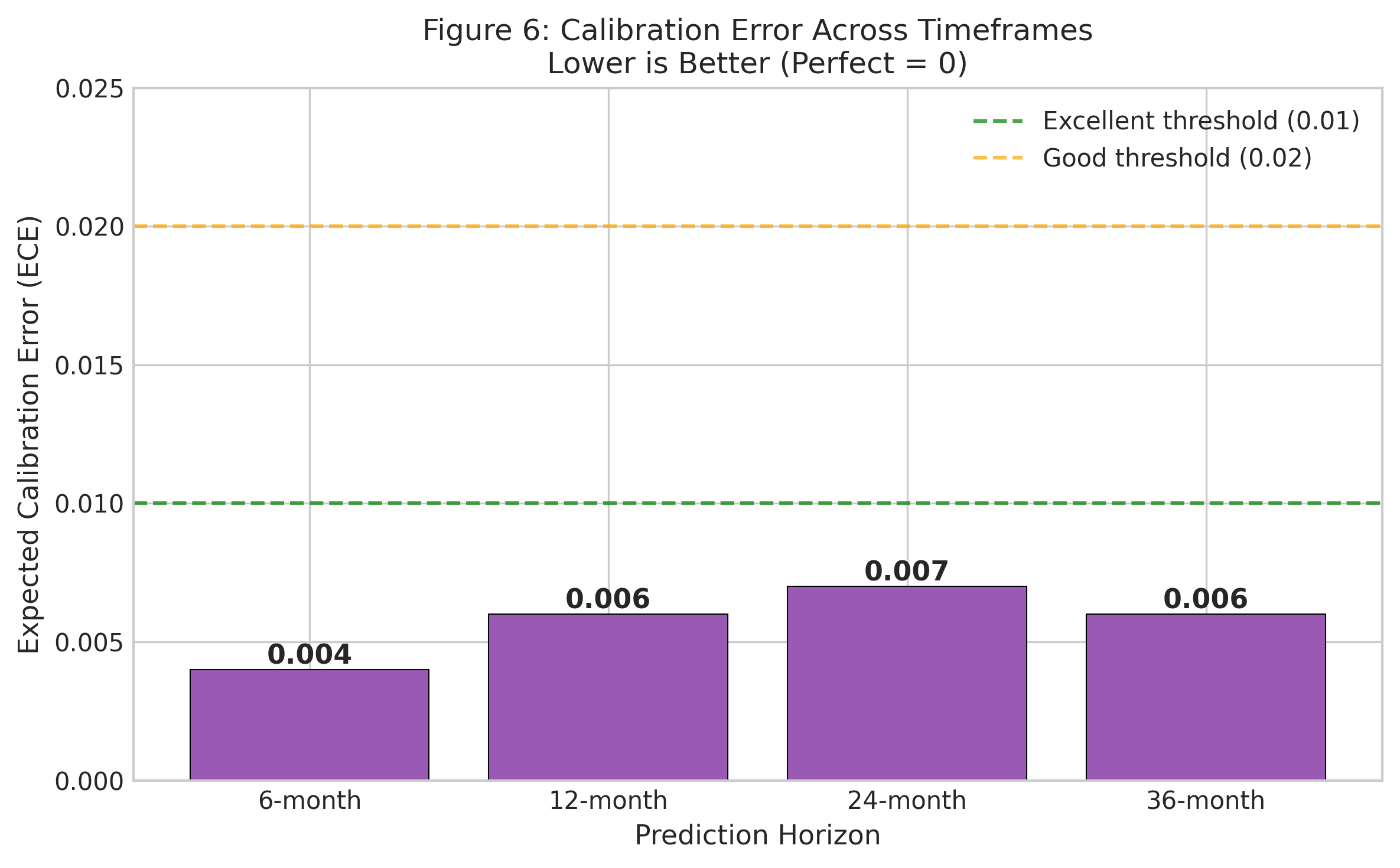

Note: Expected Calibration Error (ECE) remained below 0.01 across all prediction timeframes, indicating excellent calibration where predicted probabilities closely matched observed event rates in the synthetic validation cohort. ECE was computed as the weighted average of |predicted probability - observed proportion| across decile bins. Calibration achieved on synthetic data may not transfer to real populations with different base rates and covariate distributions.

#### Supplementary Figure S3

**Predicted Risk Distributions Across Timeframes**

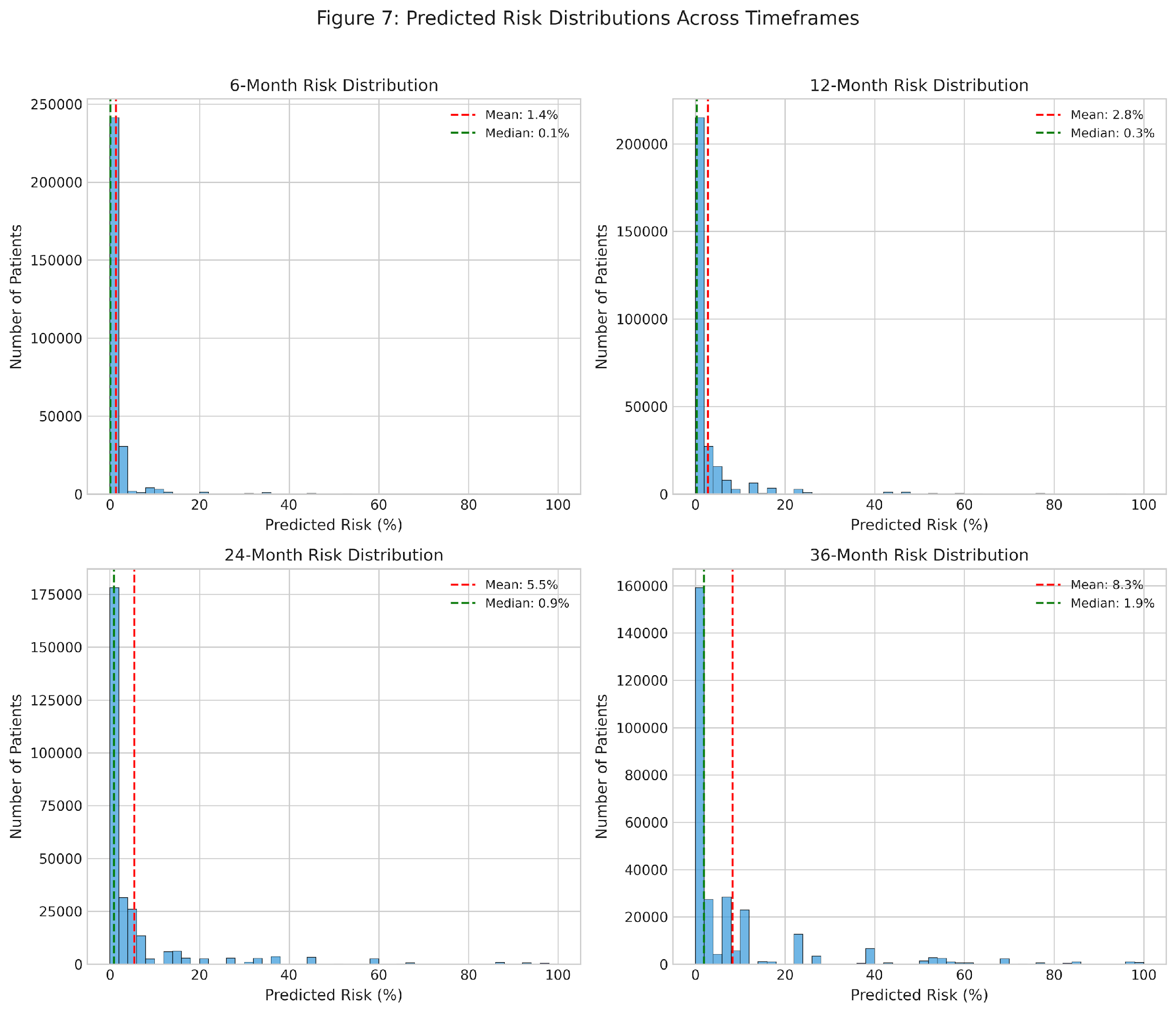

Note: Predicted risk distributions show right-skewed patterns across all timeframes, with the majority of patients classified at low risk (<15%). As expected, longer prediction horizons shift distributions rightward, reflecting increased cumulative risk over time. The 36-month predictions show greater dispersion, consistent with accumulated uncertainty over longer follow-up periods. These distributions reflect the synthetic cohort composition and may differ substantially in real patient populations.

.
